# Distinct clinical phenotypes and their neuroanatomic correlates in chronic traumatic brain injury

**DOI:** 10.1101/2025.01.27.25321200

**Authors:** Raj G. Kumar, Enna Selmanovic, Natalie Gilmore, Lisa Spielman, Lucia M. Li, Jeanne M. Hoffman, Yelena G. Bodien, Samuel B. Snider, Holly J. Freeman, Nicola L. de Souza, Christine L. Mac Donald, Brian L. Edlow, Kristen Dams-O’Connor

**Author notes:** Corresponding author: Kristen Dams-O’Connor PhD One Gustave L. Levy Place, Box 1163 New York, NY 10029 Email: kristen.dams-o’.

## Abstract

Accumulating evidence of heterogeneous long-term outcomes after traumatic brain injury (TBI) has challenged longstanding approaches to TBI outcome classification that are largely based on global functioning. A lack of studies with clinical and biomarker data from individuals living with chronic (>1 year post-injury) TBI has precluded refinement of long-term outcome classification ontology. Multimodal data in well-characterized TBI cohorts is required to understand the clinical phenotypes and biological underpinnings of persistent symptoms in the chronic phase of TBI.

The present cross-sectional study leveraged data from 281 participants with chronic complicated mild-to-severe TBI in the Late Effects of Traumatic Brain Injury (LETBI) Study. Our primary objective was to develop and validate clinical phenotypes using data from 41 TBI measures spanning a comprehensive cognitive battery, motor testing, and assessments of mood, health, and functioning. We performed a 70/30% split of training (n=195) and validation (n=86) datasets and performed principal components analysis to reduce the dimensionality of data. We used Hierarchical Cluster Analysis on Principal Components with k-means consolidation to identify clusters, or phenotypes, with shared clinical features. Our secondary objective was to investigate differences in brain volume in seven cortical networks across clinical phenotypes in the subset of 168 participants with brain MRI data. We performed multivariable linear regression models adjusted for age, age-squared, sex, scanner, injury chronicity, injury severity, and training/validation set.

In the training/validation sets, we observed four phenotypes: 1) mixed cognitive and mood/behavioral deficits (11.8%; 15.1% in the training and validation set, respectively); 2) predominant cognitive deficits (20.5%; 23.3%); 3) predominant mood/behavioral deficits (27.7%; 22.1%); and 4) few deficits across domains (40%; 39.5%). The predominant cognitive deficit phenotype had lower cortical volumes in executive control, dorsal attention, limbic, default mode, and visual networks, relative to the phenotype with few deficits. The predominant mood/behavioral deficit phenotype had lower volumes in dorsal attention, limbic, and visual networks, compared to the phenotype with few deficits. Contrary to expectation, we did not detect differences in network-specific volumes between the phenotypes with mixed deficits versus few deficits.

We identified four clinical phenotypes and their neuroanatomic correlates in a well-characterized cohort of individuals with chronic TBI. TBI phenotypes defined by symptom clusters, as opposed to global functioning, could inform clinical trial stratification and treatment selection. Individuals with predominant cognitive and mood/behavioral deficits had reduced cortical volumes in specific cortical networks, providing insights into sensitive, though not specific, candidate imaging biomarkers of clinical symptom phenotypes after chronic TBI and potential targets for intervention.

## Introduction

Traumatic brain injury (TBI) is a major cause of chronic disability worldwide.^1^ The symptoms present in the first year after injury – the time period on which most TBI studies have focused^2,3^ – are not necessarily indicative of the spectrum of chronic symptoms experienced by TBI survivors after the first year. Long-term TBI clinical sequela can include a range of evolving cognitive, mood, behavioral, and physical challenges, which do not manifest uniformly in survivors.^4–7^

TBI outcomes have historically been classified based on oversimplified indices of global function^8,9^ that have an opaque link to pathology,^9^ and are not actionable for guiding treatment. Most Phase III TBI clinical trials^10,11^ have classified outcome based on the Glasgow Outcome Scale-Extended (GOS-E).^7^ The GOS-E consists of eight coarse categories that map to a continuum of disability; however, this measure does not distinguish the nature of clinical symptoms that contribute to disability, thereby limiting its clinical relevance. For example, two individuals may both be classified in the “moderate disability” category on the GOS-E despite having a completely distinct constellation of symptoms (e.g., predominantly cognitive versus physical symptoms) attributable to different underlying pathologies that warrant entirely different treatment approaches.^12–14^ Moreover, studies frequently dichotomize the GOS-E.^15^ While a binary outcome (e.g., favorable and unfavorable recovery) is concise and intuitive, it further compromises measurement precision on an already coarse scale.^16^ Indeed, the lack of a granular outcome classification system has been posited by experts to be a key reason for the failure of many Phase III clinical trials to demonstrate the efficacy of novel therapies.^17,18^

While most prior TBI clinical trials have focused on acute treatments, there is growing priority^19^ to advance scientific understanding of the long-term sequelae of TBI and potential intersections with other health conditions including dementia. As such, classification of patients with TBI based on chronic care needs is an important area for research; few TBI studies to date have both clinical and biomarker data more than a year post injury. Clinical trials of novel pharmacological and non-pharmacological treatments for chronic TBI patients requires a paradigm shift away from conventional approaches that base study eligibility on distally-assessed acute TBI severity, in favor of criteria based on patients’ enduring clinical symptoms and contemporaneous neuroanatomic features.

In recent years, international efforts to establish more sophisticated approaches to TBI characterization reflect a growing interest in TBI “phenotyping”.^20^ Studies have used a variety of methodological approaches, including model-based (e.g., latent class modeling^21^), supervised machine leaning (e.g., classification and regression trees^22^), and unsupervised machine learning (e.g., cluster analyses^23,24^). While these methods have different assumptions and mathematical formulae, their purpose is similar: to identify subgroups of individuals who share similar profiles of performance-based and/or self-reported clinical data. Notably, most studies of this type have focused on characterizing subacute post-concussion or mild TBI symptom profiles within the first year post-injury; it is unclear whether phenotypes differ into the chronic TBI period. Similar patterns of clinical symptom subtypes have emerged from past work, including phenotypes with mental health/behavioral symptoms and phenotypes with somatic/functional symptoms.^20^ Few phenotyping studies have investigated clinically accessible biomarker correlates; many also exclude those with large cortical lesions that interrupt neuroimaging processing pipelines, which limits their generalizability and precludes elucidation of pathophysiological and anatomical features that may inform intervention targets.^25,26^ To address the aforementioned knowledge gaps, we used unsupervised machine learning methods agnostic to any existing TBI classification to analyze our high-dimensional data with fewer assumptions. We then investigated neuroimaging-based biomarkers of identified clinical phenotypes. We implemented an extensive pre-processing pipeline that addresses potential sources of bias from past studies, including imputation for non-random missingness of clinical data. We also used a novel lesion-correction methodology to avoid excluding participants with large lesions from neuroimaging analyses.

The primary objective of the present study builds upon the emerging TBI phenotyping literature by leveraging multimodal data from the Late Effects of TBI (LETBI) Study to identify chronic TBI clinical phenotypic clusters. The LETBI study collects an extensive battery of clinical symptom assessments and performance-based tests of cognitive and motor function, in addition to neuroimaging, in a sample of individuals who are at least one-year post TBI. Our secondary objective is to address a key gap in the literature by evaluating associations between the identified clinical phenotypes and network-based cortical neuroimaging biomarkers to describe the neuroanatomic correlates of clinical phenotypes.

## Materials and Methods

### Participants

The present study leveraged research registries and community-based outreach to enroll 281 participants in the multi-center LETBI study at the University of Washington (UW) and the Icahn School of Medicine at Mount Sinai (ISMMS).^27^ Participants were eligible if they met the following criteria: age ≥ 18 years, English speaking, complicated mild^28^, moderate or severe TBI^29^ and at > 1-year post-injury. The LETBI study inclusion criteria and recruitment methods differ in important ways from other TBI cohorts because participants were not enrolled solely from a clinic or hospital, which allows inclusion of individuals who did not immediately seek care for their TBI. The parent LETBI study is a longitudinal prospective cohort study; however, for the present analysis we used cross-sectional data acquired at the first LETBI study visit. Data for this study were collected between June 2014 and August 2022. Local institutional review boards approved this protocol at both study sites and written informed consent was provided by participants or proxy decision-makers. The derivation of the analytic sample for our primary and secondary objectives are in Fig 1.

**Fig 1:**
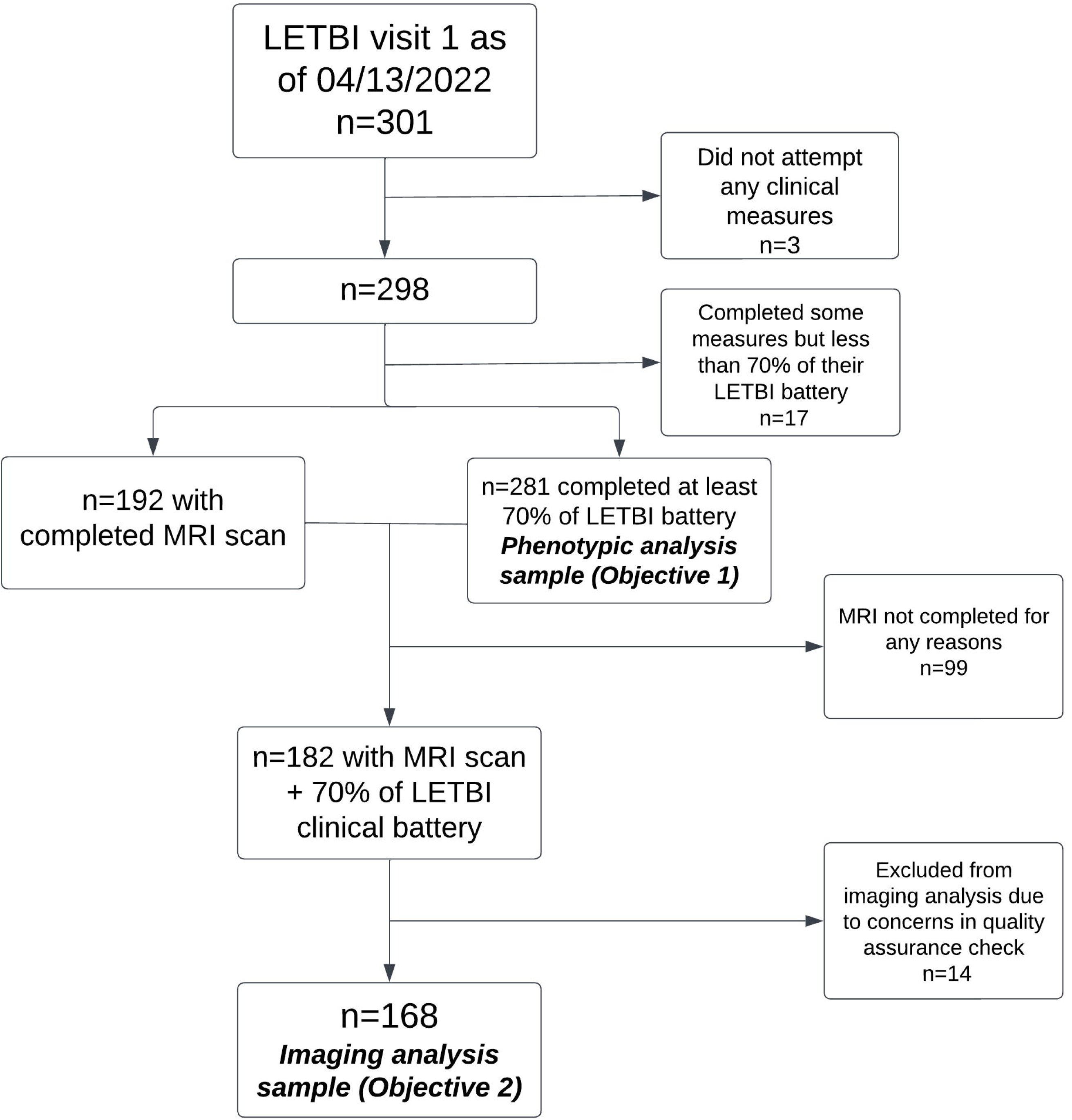
Study Flow Diagram. We provide details on the derivation of the phenotypic (primary objective) and neuroimaging analytic (secondary objective) samples.

### Demographic and Injury Characteristics

Demographic data included age, sex, education (less college degree, college degree or higher), race (White, Black, other), Hispanic ethnicity. Pre-injury characteristics included: marital status (never married, married/partnered, divorced/widowed, and employment status (working/student, unemployed and looking for work, retired, disabled, other). Information on TBI injury history was collected using the Brain Injury Screening Questionnaire (BISQ)^30^, a semi-structured measure of retrospective TBI ascertainment that relies on cues to query lifetime head injury, repetitive head impact, and TBI exposure. Using the BISQ, we derived the following calculated variables of lifetime TBI history in this study: years since most recent TBI, years since first TBI, and injury severity based on the most severe lifetime injury. Our approach for injury severity characterization was based on presence and duration of loss of consciousness and/or post-traumatic amnesia. Specifically, we defined the mild/moderate/severe designation as defined by the American Congress of Rehabilitation Medicine^31^ and the Department of Defense.^32^

### Clinical Measures

The LETBI study collects data using 41 distinct clinical measures. The assessment battery consists of combination of performance-based and patient-reported outcomes, including a comprehensive neuropsychological battery, objective physical and motor testing, and self-report assessments pertaining to mood, health and functioning. The neuropsychological battery spanned the following domains and associated tests: learning and memory (Wechsler Memory Scale® (WMS) Fourth Edition^33^ Logical Memory: I and II; Rey Complex Figure Test^34^: Immediate, Delayed, Recognition; California Verbal Learning Test^35^ (CVLT): Immediate Recall 1-5, Long Delay and Short Delay Free Recall), verbal fluency (Controlled Oral Word Association Test^36^ (COWAT) Animals and Words), processing speed (Trails A and B^37^; Wechsler Adult Intelligence Scale^38^® (WAIS), Fifth Edition: Coding and Symbol Search), attention/working memory (WAIS Digit Span), and executive functioning (CVLT: Semantic and Serial Clustering). The mood measures included self-report questionnaires on anxiety (Quality of Life in Neurological Disorders^39^ (Neuro-QOL): Anxiety), depression (Neuro-QOL: Depression), fatigue (Neuro-QOL: Fatigue; RAND Short Form 36 item^40^ (SF-36) Energy/Fatigue), social functioning (*Neuro-QOL*: Social; RAND SF-36 Social Functioning), satisfaction with life (Satisfaction with Life Scale), and impulsivity and aggression (Barratt Impulsivity Scale^41^® Second Edition: Non-planning, Motor, and Attention). Measures of health and function included assessments of independence with activities of daily living (RAND SF-36 Physical Functioning; RAND SF-36 Physical Limitations), motor performance (Unified Parkinson’s Disease Rating Scale^42^ (UPDRS) Part III Motor Exam; Dominant Hand Grip Strength), self-rated health and self-rated memory (based on 5-point Likert rating scale items from Midlife in the United States (MIDUS) Study), alcohol and substance abuse (Alcohol, Smoking and Substance Involvement Screening Test (ASSIST): Illicit drug, Tobacco, and Alcohol use subscales), and chronic pain (RAND SF-36 pain subscale). We included individuals in the present study if they completed at least 70% of their clinical assessment, which led to inclusion of 281 of a possible 298 participants (94.3%). The 70% minimum criteria was based on having sufficient data for missing data imputation of clinical measures, which is discussed in further detail below.

### Neuroimaging Data Acquisition

For our secondary aim, a subset of study participants (n=168) were examined who had completed T1-weighted multi-echo magnetization-prepared rapid gradient echo (MEMPRAGE) imaging at either ISMMS or UW.^27^ At ISMMS, we acquired the T1 MEMPRAGE sequence from a 3T Siemens Skyra (Siemens Medical Solutions, Erlangen, Germany) scanner with a 32-channel head coil. At UW, we acquired the T1 MEMPRAGE sequence from either a 3T Philips Achieva (n = 59) or 3T Ingenia Elition (n = 58) MRI scanner with a 32-channel head coil. All sequences were 1 mm isotropic spatial resolution. We harmonized acquisition sequences across both sites as previously described^27^ with only slight differences in the echo time x 4 = 1.67 ms/3.47ms/5.27ms/7.07ms and inversion time = 1100 ms for UW relative to ISMMS. To avoid confounding, we included scanner type as a covariate in the regression analyses.

### Neuroimaging Data Processing

We processed T1-weighted MEMPRAGE data using the “recon-all” pipeline in Freesurfer^43^ version 7.2.0, which involves extraction of cortical surfaces, parcellation of cortical regions, and segmentation of subcortical structures using the Desikan-Killiany atlas. We mapped the 1,000-node parcellation of the Yeo-7 network atlas^44^ to each participant’s surface. We then projected atlas labels from surface vertices into the T1 anatomical volume for each participant. We extracted cortical volume estimates for parcellations within each of the Yeo-7 networks^45^: visual, somatomotor, dorsal attention, salience (i.e., ventral attention), limbic, executive control, and default mode. We summed volume estimates within the nodes of each network to obtain the total volume for each network.

Cortical lesions can lead to inaccurate surface rendering in automated segmentation pipelines, such as FreeSurfer, resulting in imprecise or “failed” segmentations and ultimately excluding participants from studies.^46^ Exclusion of participants with large cortical lesions from prior TBI studies^25,26^ can contribute to non-random missingness bias and limits the generalizability of findings; we applied a lesion-correction methodology to overcome this limitation. Specifically, our team (H.J.F., N.G., and S.B.S.) screened all T1-weighted images for the presence of cortical lesions. Lesions that disrupted the cerebral cortex were traced by a study investigator (H.J.F.) using the Voxel Edit tool in FreeView, and multiple investigators (H.J.F., N.G., S.B.S.) confirmed that cortical lesion tracings covered the entire lesioned area and that the lesion boundary did not extend into ventricles. Cortical lesions could extend into surrounding white matter. However, we did not trace lesions that were isolated to white matter given that these lesions are difficult to localize to specific networks.

We then merged cortical lesions with the initial white matter segmentation, generating a “lesion-corrected” white matter mask. The initial white matter segmentation generated by Freesurfer software’s recon-all serves as a reference for surface placement later in the processing pipeline. However, disruptions in the white matter segmentation caused by lesions can result in misaligned surfaces that incorrectly pass through cortical regions affected by the injury. By merging the lesion segmentation with the white matter segmentation, these gaps are bridged, promoting more accurate surface placement in relation to the cortex. Using this lesion-corrected white matter mask, we re-executed “recon-all”, resulting in corrected surfaces for the pia and grey-white matter boundary that included the lesioned areas. We then registered the Yeo 7-network atlas^45^ via surface registration for each participant, and each network was projected into the T1 anatomical volume space. Finally, lesioned areas within cortical network labels were assigned a volume of zero, allowing us to quantitatively account for a lesion’s impact on the overall structural integrity of each network. Partial lesion overlap affects only the portion of a region that intersects with the lesion label. To illustrate an exemplar of the implementation of our novel lesion correction methodology in practice, we have provided “before” and “after” MRI scans in Supplementary Fig 1.

### Neuroimaging Data Quality Assessments

We assessed FreeSurfer segmentations for all participants for completeness and accuracy (i.e., precision of cortical and subcortical segmentation), with special attention to areas known to exhibit segmentation errors (e.g., thalamic and temporal regions). We adapted our scale to rate segmentation quality from a previously published quality assessment scale^46^ that is described in the Supplementary Table 1. We excluded a total of 20 participants from analysis after visual inspection revealed major disruptions in pial and white surfaces and/or exclusion or mislabeling of cortical areas (n=9), or due to failure to reconstruct surfaces after applying the lesion correction (n=11).

### Statistical Analysis

To illustrate the complex, multi-step nature of our analysis pipeline, we have summarized our pre-processing and analysis steps in Fig 2.

**Fig 2:**
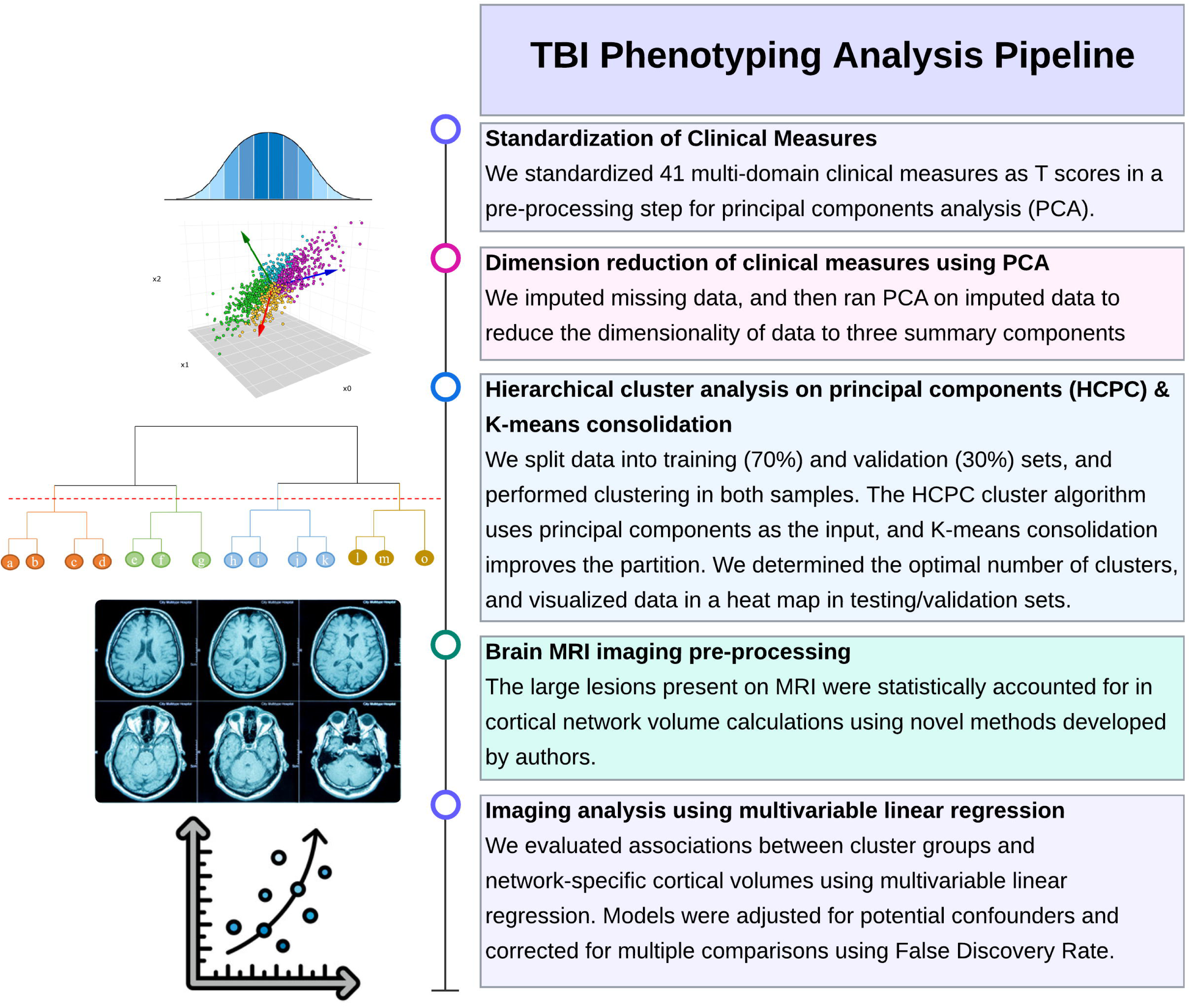
TBI Phenotyping Analysis Pipeline. Includes pre-processing steps for clinical and neuroimaging MRI data. Last two steps were among the subgroup with MRI data. Visuals in the left column are not based on data from the present study but meant as theoretical depictions.

#### Clinical phenotyping analyses

We used unsupervised machine learning methods to identify clusters of participants with similar profiles as defined by clinical data across multiple dimensions (e.g., cognitive, mood, motor). To accomplish this, we leveraged a three-tier pipeline of common multivariate approaches: principal components analysis (PCA), hierarchical clustering, and k-means partitioning clustering implemented using the R package *FactoMineR* as described below.^47^ Cluster results were determined in an initial training set (random 70% of the sample; n=195), and then validated in an internal validation set (random 30% of the sample; n=86).

First, we normalized all continuous data elements against reference populations to be on the same standardized scale. Next, we investigated the missingness of data. Most statistical software packages that implement PCA require a complete case analysis, relying on a highly tenuous missing completely at random (MCAR) assumption for valid results. This assumption is often impractical in clinical TBI studies, where it is common for participants with impairments to selectively complete tests, particularly for long research interviews with multidimensional data. In this study, we handled missing data using a regularized iterative pre-processing PCA algorithm.^48^ Briefly, this method starts with an initial placeholder value of single mean imputation calculated from observed values. Then a PCA is iteratively performed on the whole dataset to re-estimate missing data parameters until achieving convergence according to an expectation maximization algorithm.^49^ In short, we used observed data (summarized through principal dimensions) as predictors to model missing data parameters. Importantly, this method assumes a more relaxed Missing at Random (MAR) assumption (i.e., missing data is random conditional on observed (non-missing) variables, represented mathematically herein in the form of principal components). Prior simulation studies found that this method outperforms other methods to handle missing data in the setting of high-dimensional data analyses where alternative methods—namely multiple imputation—can sometimes be unwieldy to interpret.^50^ The missing data imputation was implemented using the *missMDA* package in R.^51^ As stated above, we included participants with at least 70% of their LETBI interview completed, so no more than 30% of data was imputed for a given participant.

Using our imputed dataset, we then performed standard PCA on our 41 data elements spanning multiple domains to reduce the dimensionality and noise into a small set of components. Our decision of number of principal components was based on inspection of the Scree plot and the associated tradeoff of maximizing variance explained and minimizing the noise. Our principal components were unrotated because we were not directly interpreting the PC output, rather it served as a dimension reduction intermediary step before the cluster analysis (described below).

Next, we randomly divided our sample into a 70% (n=195) training and 30% (n=86) validation set, to implement and validate the hierarchical cluster analysis on principal components (HCPC). Hierarchical cluster analysis is an agglomerative “bottom-up” method where each individual starts as their own cluster, and then similar pairs of clusters are grouped together based on their multivariate data (i.e., represented using principal component scores). This process is iterated hierarchically until all individuals are “related” to all other individuals. Relative similarity is based on Euclidean distances, and we used Ward’s minimum variance method which minimizes between cluster differences in variance.^52^ We used this clustering method – which uses selected principal component scores as the input variables – for our research question because we included 41 different clinical measures that would have been computationally intensive to determine meaningful clusters.

The primary decision point for cluster analyses is determining the most appropriate number of clusters, for which best practice dictates a balance between evaluating quantitative fit indices and assessing clinical interpretability. Using the training set, an initial cut point was automatically chosen on the dendrogram (a tree-like hierarchical visual representation of the relationship between individuals) based on minimizing within-cluster inertia (i.e., distance from individual points to cluster centroid). We further improved the initial HCPC partition through an additional k-means consolidation step. This doubly robust approach merges elements of hierarchical and partitioning methods, increasing accuracy and precision of cluster assignment compared to using either method alone.^53^ Furthermore, we quantitatively and visually evaluated cluster metrics using the Dunn Index^54^, Connectivity^55^, Silhouette Index^56^ for multiple different specifications of numbers of clusters calculated using the *clValid* package in R.^57^ Based on converging evidence from the training set, the presumed number of selected clusters was tested in the (independent) validation set to assess the internal reliability and validity of the clusters.

To illustrate the multivariate characteristics of each cluster, we created a heat map of the 41 input measures by cluster using the *pheatmap* R package.^58^ To aid in interpretability of the heat map visualization, we transformed measures where higher scores indicated better performance, such that high values across all measures corresponded to poorer performance. We also described sample characteristics by cluster assignment to understand the composition of individuals in each cluster.

#### Cortical volumetric analyses

For our secondary objective, we evaluated associations between cluster membership and the total volume of each of the Yeo 7-networks^45^ using a series of multivariable linear regression models for each network. Because our MRI sample for the secondary objective was reduced relative the sample from the first objective, we checked for the presence of selection bias. First, we descriptively compared characteristics of participants who were included in the first and secondary objective versus those participants who were in first objective-only (i.e., without MRI). Next, as a sensitivity analysis to ensure the consistency of our cluster results in the smaller sample, we replicated the primary cluster analyses among only participants in the secondary objective sample. To have a sufficiently powered sample for neuroimaging models in our smaller sample, we combined the training and validation sets for these models, and adjusted for an indicator of sample set. All models were adjusted for age, age-squared, sex, scanner type, time since most recent injury, severity of most severe injury (moderate/severe TBI versus mild TBI), and training/testing set, consistent with confounders considered in recent TBI neuroimaging studies.^59,60^ We accounted for multiple comparisons in network-specific neuroimaging models using a False Discovery Rate (FDR) approach.^61^ We plotted unadjusted cortical volumes by cluster, and calculated model-based estimated marginal means using a least-squares mean approach^62^ to facilitate a comparison of mean volumes across all four clusters, adjusted for covariates. We further conducted post-hoc pairwise statistical comparisons of least square mean differences in cortical network for all clinical phenotype contrasts.

## Results

### Sample characteristics

We provide the demographic and clinical characteristics of the total sample by training and validation set in Table 1. Briefly, the average age at interview of the total sample was 57.5 (SD=16), and 34.2% percent were female, 86.1% were White, 8.2% were Hispanic, and 72.6% had at least a college degree. Approximately one-third of the sample was currently working or a student, while approximately 30% and 24% reported being retired or not working because of a disability, respectively. The average time since the most recent TBI and first lifetime TBI was 8.2 years and 27.5 years, respectively. A majority of the sample had at least one severe TBI (59.4%) in their lifetime. There were more females with slightly less time since their most recent injuries in the training set, otherwise there were no meaningful differences on any variables between the training and validation sets.

**Table 1:**
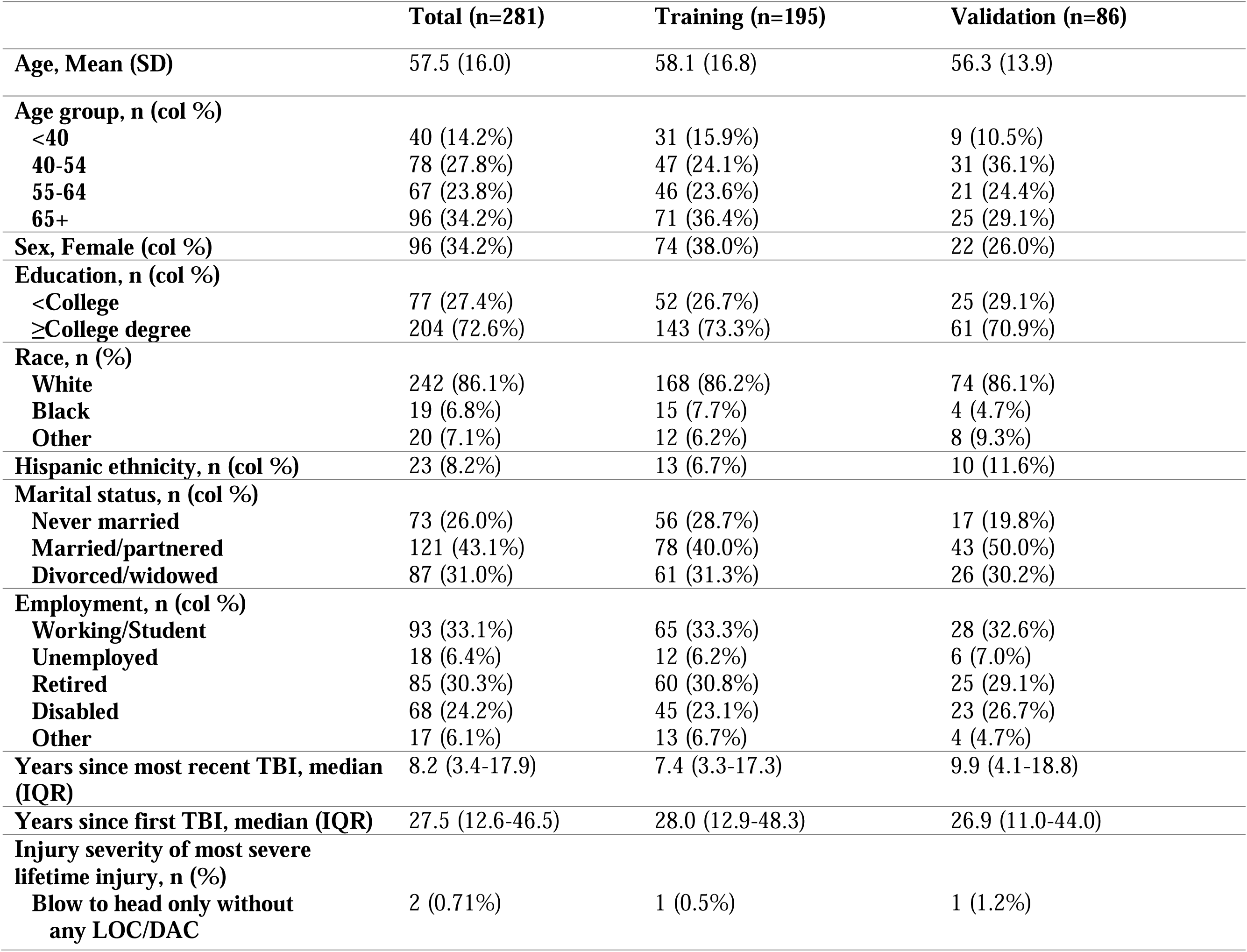
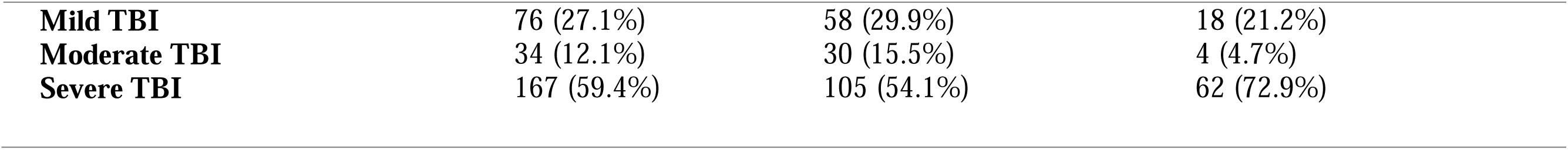
Characteristics of the sample by training and validation set.

|  | **Total (n=281)** | **Training (n=195)** | **Validation (n=86)** |
| --- | --- | --- | --- |
| **Age, Mean (SD)** | 57.5 (16.0) | 58.1 (16.8) | 56.3 (13.9) |
| **Age group, n (col %)** |  |  |  |
| <40 | 40 (14.2%) | 31 (15.9%) | 9 (10.5%) |
| 40-54 | 78 (27.8%) | 47 (24.1%) | 31 (36.1%) |
| 55-64 | 67 (23.8%) | 46 (23.6%) | 21 (24.4%) |
| 65+ | 96 (34.2%) | 71 (36.4%) | 25 (29.1%) |
| **Sex, Female (col %)** | 96 (34.2%) | 74 (38.0%) | 22 (26.0%) |
| **Education, n (col %)** |  |  |  |
| <College | 77 (27.4%) | 52 (26.7%) | 25 (29.1%) |
| ≥College degree | 204 (72.6%) | 143 (73.3%) | 61 (70.9%) |
| **Race, n (%)** |  |  |  |
| White | 242 (86.1%) | 168 (86.2%) | 74 (86.1%) |
| Black | 19 (6.8%) | 15 (7.7%) | 4 (4.7%) |
| Other | 20 (7.1%) | 12 (6.2%) | 8 (9.3%) |
| **Hispanic ethnicity, n (col %)** | 23 (8.2%) | 13 (6.7%) | 10 (11.6%) |
| **Marital status, n (col %)** |  |  |  |
| Never married | 73 (26.0%) | 56 (28.7%) | 17 (19.8%) |
| Married/partnered | 121 (43.1%) | 78 (40.0%) | 43 (50.0%) |
| Divorced/widowed | 87 (31.0%) | 61 (31.3%) | 26 (30.2%) |
| **Employment, n (col %)** |  |  |  |
| Working/Student | 93 (33.1%) | 65 (33.3%) | 28 (32.6%) |
| Unemployed | 18 (6.4%) | 12 (6.2%) | 6 (7.0%) |
| Retired | 85 (30.3%) | 60 (30.8%) | 25 (29.1%) |
| Disabled | 68 (24.2%) | 45 (23.1%) | 23 (26.7%) |
| Other | 17 (6.1%) | 13 (6.7%) | 4 (4.7%) |
| **Years since most recent TBI, median (IQR)** | 8.2 (3.4-17.9) | 7.4 (3.3-17.3) | 9.9 (4.1-18.8) |
| **Years since first TBI, median (IQR)** | 27.5 (12.6-46.5) | 28.0 (12.9-48.3) | 26.9 (11.0-44.0) |
| **Injury severity of most severe lifetime injury, n (%)** |  |  |  |
| Blow to head only without any LOC/DAC | 2 (0.71%) | 1 (0.5%) | 1 (1.2%) |
| **Mild TBI** | 76 (27.1%) | 58 (29.9%) | 18 (21.2%) |
| **Moderate TBI** | 34 (12.1%) | 30 (15.5%) | 4 (4.7%) |
| **Severe TBI** | 167 (59.4%) | 105 (54.1%) | 62 (72.9%) |

### Dimension reduction of multivariate data using PCA and cluster analysis

We standardized 41 measures spanning multiple domains of cognitive, motor, physical and mental health, and imputed missing data using the procedures detailed above. In the imputed dataset, we reduced the dimensionality of the data into three principal components that represented 45.2% of the variance in the data. We have presented the top clinical measures that loaded to each of the first three principal components in Supplementary Table 2.

We determined the optimal number of hierarchical clusters was a four-cluster assignment based on quantitative evidence from cluster validity indices (Supplementary Table 3), in addition to visual evidence from the dendrogram and cluster Scree plot (Supplementary Fig 2). We also visually inspected internal validity and reliability by comparing the stability in the centroid coordinates between the training (Figure 3) and validation (Supplementary Fig 3) sets. We determined that, compared the four-cluster assignment, the three-cluster assignment was more unreliable and inconsistent in internal validation (Supplementary Fig 4).

**Fig 3:**
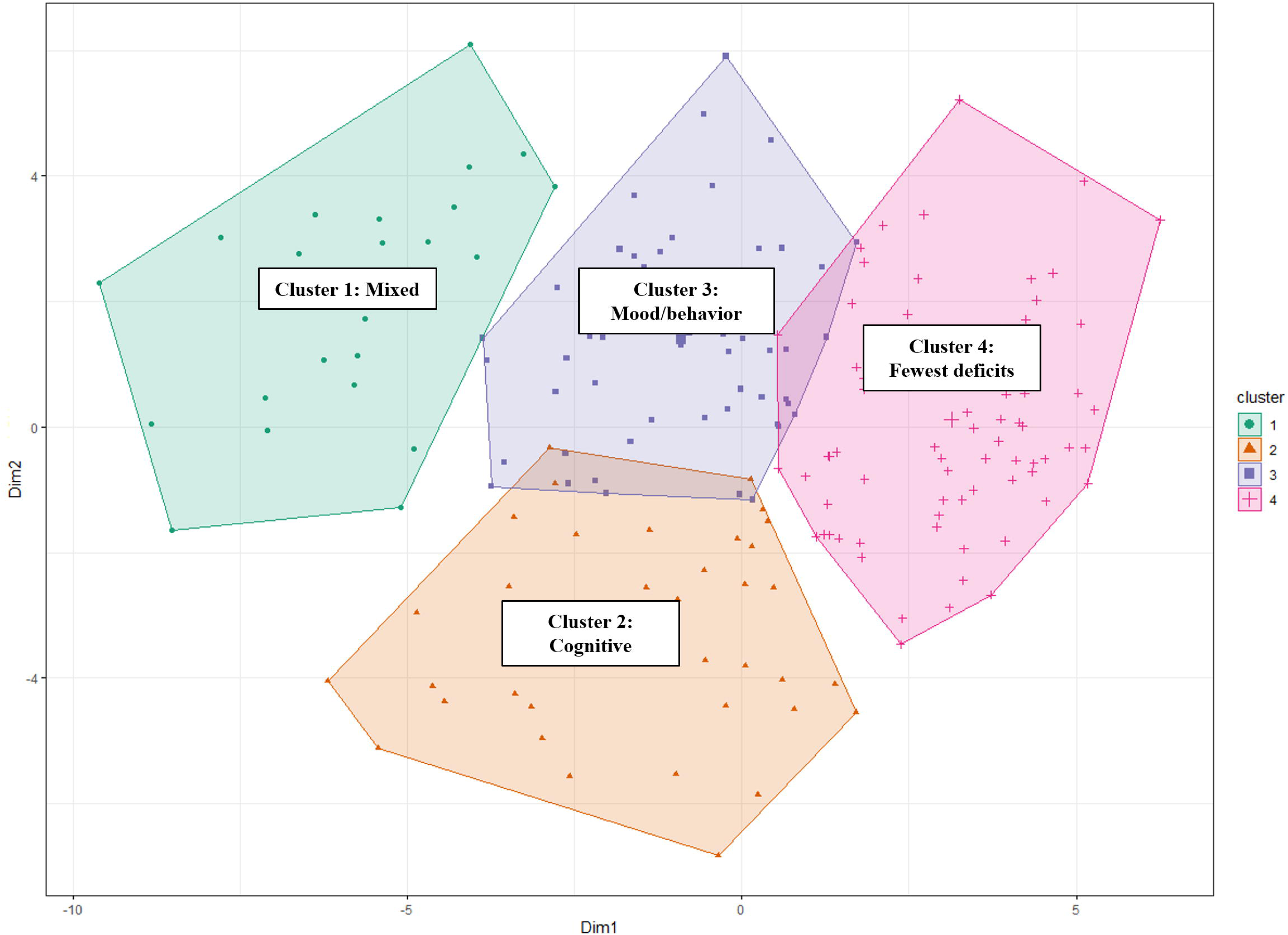
Hierarchical cluster group assignment (training set). Hierarchical cluster group assignment in the training set. The results are based on a Hierarchical Cluster on Principal Components (HCPC). Here, each participant in the training sample is depicted in the x-y coordinate space based on their PC1 vs. PC2 scores. The cluster membership of each participant is color coded.

### Heat map: Multivariate illustration by cluster/phenotype

Using the four-cluster assignment, we generated a heat map of the average values by measure in the training (Fig 4) and validation (Supplementary Fig 5) sets. We observed a similar clustering pattern for the four clusters in the training and validation sets. Individuals in cluster 1 (11.8% and 15.1% of training and validation samples, respectively) had mixed trait (referred hereafter as Cluster 1_mixed_) deficits spanning across most multidimensional clinical measures. Persons in cluster 2 (training: 20.5% and validation: 23.3%) had predominant cognitive deficits (referred hereafter as Cluster 2_cognitive_), while those in cluster 3 (training: 27.7% and validation: 22.1%) predominantly had mood and behavioral deficits (referred hereafter as Cluster 3_mood/behavior_). Individuals in cluster 4 (training: 40% and validation: 39.5%) had the fewest cognitive, mood, neurobehavioral, and physical deficits in the sample (referred hereafter as Cluster 4_fewest deficits_).

**Fig 4:**
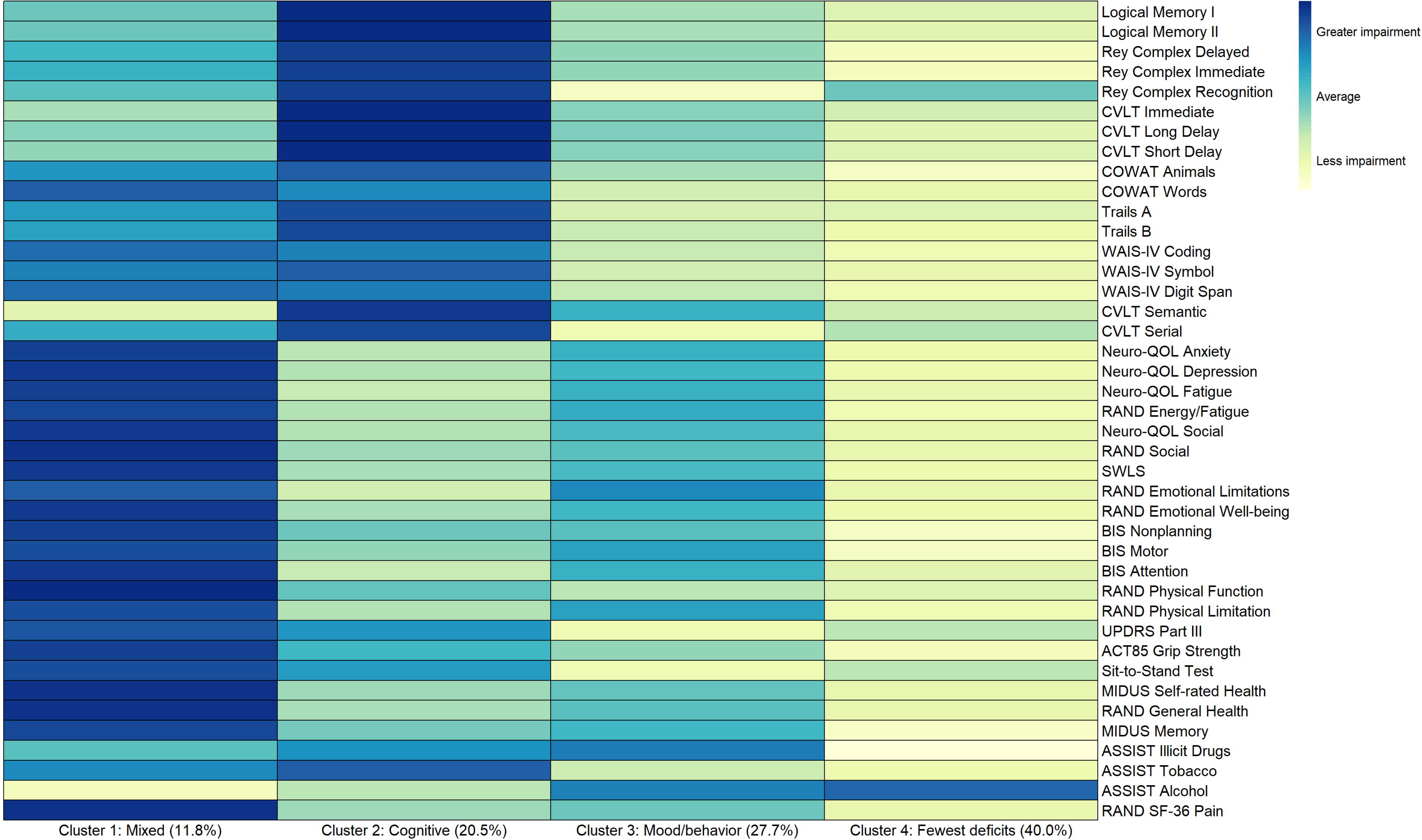
Heat map characterizing average values of neurobehavioral measures by cluster assignment (training set; n=195). Heat map characterizing average values of neurobehavioral measures by cluster assignment in the training set. The measures have all been transformed such that darker colors represent greater impairment, and lighter colors represent less impairment. Based on the findings, we have assigned the following qualitative descriptors of each cluster: Cluster 1: Mixed deficits; Cluster 2) Predominant cognitive deficits; Cluster 3: Predominant mood and behavioral deficits; Cluster 4: Relatively few deficits.

### Characteristics of the sample by cluster

In Table 2, we have descriptively characterized the four clusters. Individuals in Cluster 4_fewest deficits_ were on average older, more educated, and more chronically removed from their most recent injury (i.e., average 10.1 years post-injury). Individuals in Cluster 3_mood/behavior_ were on average the youngest cluster, and were well educated. Other noteworthy findings were that females were less likely than males to be in Cluster 4_fewest deficits_, and individuals with less than a college degree disproportionately tended to belonged to Cluster 2_cognitive_. The Cluster 1_mixed_ group more often had severe TBI and were more recently injured (i.e., average 5.6 years post-injury). When we examined current employment status by cluster, we found that 35.6% and 42.9% of those who were working or currently a student belonged to Cluster 3_mood/behavior_ or Cluster 4_fewest deficits_ groups, respectively. Persons who were unemployed, but looking for work, were most often in Cluster 2_cognitive_, and those who reported not working because of a current disability were spread mostly evenly between clusters 1-3, with less than 10% belonging to Cluster 4_fewest deficits_.

**Table 2:** Descriptive characterization of the clusters (full sample)

|  | **Cluster 1**<br>*Mixed deficits* | **Cluster 2**<br>*Predominant cognitive deficits* | **Cluster 3**<br>*Predominant mood/behavioral deficits* | **Cluster 4**<br>*Fewest deficits* |
| --- | --- | --- | --- | --- |
| **Row n (%)** | 36 (12.8%) | 60 (21.4%) | 73 (26.0%) | 112 (39.9%) |
| **Age at interview, Mean (SD)** | 54.7 (12.4) | 54.2 (16.0) | 51.6 (15.6) | 64.2 (14.9) |
| **Age at interview group, n (col %)** |  |  |  |  |
| **<40** | 3 (8.3%) | 11 (18.3%) | 19 (26.0%) | 7 (6.3%) |
| **40-54** | 14 (38.9%) | 20 (33.3%) | 23 (31.5%) | 21 (18.8%) |
| **55-64** | 13 (36.1%) | 18 (30.0%) | 14 (19.2%) | 22 (19.6%) |
| **65+** | 6 (16.7%) | 11 (18.3%) | 17 (23.3%) | 62 (55.4%) |
| **Sex, n (col %)** |  |  |  |  |
| **Male** | 21 (58.3%) | 37 (61.7%) | 48 (65.8%) | 79 (70.5%) |
| **Female** | 15 (41.7%) | 23 (38.3%) | 25 (34.3%) | 33 (29.5%) |
| **Education, n (col %)** |  |  |  |  |
| **Less than college degree** | 13 (36.1%) | 27 (45.0%) | 18 (24.7%) | 19 (17.0%) |
| **College degree or higher** | 23 (63.9%) | 33 (55.0%) | 55 (75.3%) | 93 (83.0%) |
| **Race, n (row %)** |  |  |  |  |
| **White** | 29 (80.6%) | 46 (76.7%) | 65 (89.0%) | 102 (91.1%) |
| **Black** | 5 (13.9%) | 7 (11.7%) | 4 (5.5%) | 3 (2.7%) |
| **Other** | 2 (5.6%) | 7 (11.7%) | 4 (5.5%) | 7 (6.3%) |
| **Hispanic ethnicity, n (col %)** | 4 (11.1%) | 8 (13.3%) | 4 (5.5%) | 7 (6.3%) |
| **Marital status, n (col %)** |  |  |  |  |
| **Never married** | 13 (36.1%) | 26 (43.3%) | 21 (28.8%) | 13 (11.6%) |
| **Married/partnered** | 11 (30.6%) | 20 (33.3%) | 30 (41.1%) | 60 (53.6%) |
| **Divorced/widowed** | 12 (33.3%) | 14 (23.3%) | 22 (30.1%) | 39 (34.8%) |
| **Employment, n (col %)** |  |  |  |  |
| **Working/Student** | 5 (13.9%) | 14 (23.3%) | 26 (35.6%) | 48 (42.9%) |
| **Unemployed, looking for work** | 2 (5.6%) | 8 (13.3%) | 3 (4.1%) | 5 (4.5%) |
| **Retired** | 6 (16.7%) | 11 (18.3%) | 15 (20.6%) | 53 (47.3%) |
| **Disabled** | 23 (63.9%) | 20 (33.3%) | 19 (26.0%) | 6 (5.4%) |
| **Other** | 0 (0%) | 7 (11.7%) | 10 (13.7%) | 0 (0%) |
| **Years since most recent TBI, median (IQR)** | 5.6 (2.9-13.2) | 6.9 (2.8-17.4) | 7.8 (3.9-13.8) | 10.1 (3.8-21.2) |
| **Years since first TBI, median (IQR)** | 22.8 (6.4-46.5) | 19.7 (7.9-36.2) | 26.9 (13.6-41.4) | 35.6 (16.4-56.0) |
| **Injury severity of most severe lifetime injury, n (col %)** |  |  |  |  |
| **Blow to head only without any LOC/DAC** | 0 (0%) | 0 (0%) | 1 (1.4%) | 1 (0.9%) |
| **Mild TBI** | 4 (11.1%) | 19 (32.2%) | 17 (23.3%) | 36 (32.4%) |
| **Moderate TBI** | 7 (19.4%) | 3 (5.1%) | 9 (12.3%) | 15 (13.5%) |
| **Severe TBI** | 25 (69.4%) | 37 (62.7%) | 46 (63.0%) | 59 (53.2%) |

**Table 3:**
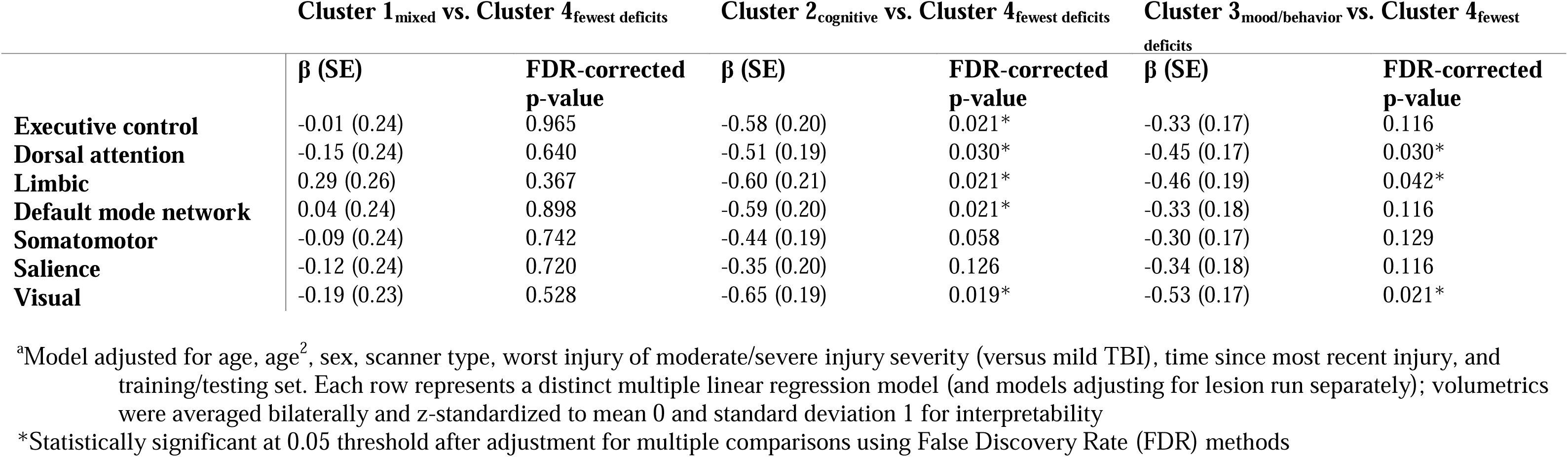
Multiple linear regression models^a^ of Yeo-7 cortex networks volumes (full sample; n=168 with concurrent neuroimaging)

|  | Cluster 1<sub>mixed</sub> vs. Cluster 4<sub>fewest deficits</sub> |  | Cluster 2<sub>cognitive</sub> vs. Cluster 4<sub>fewest deficits</sub> |  | Cluster 3<sub>mood/behavior</sub> vs. Cluster 4<sub>fewest deficits</sub> |  |
| --- | --- | --- | --- | --- | --- | --- |
| | $\beta$ (SE) | FDR-corrected p-value | $\beta$ (SE) | FDR-corrected p-value | $\beta$ (SE) | FDR-corrected p-value |
| **Executive control** | -0.01 (0.24) | 0.965 | -0.58 (0.20) | 0.021* | -0.33 (0.17) | 0.116 |
| **Dorsal attention** | -0.15 (0.24) | 0.640 | -0.51 (0.19) | 0.030* | -0.45 (0.17) | 0.030* |
| **Limbic** | 0.29 (0.26) | 0.367 | -0.60 (0.21) | 0.021* | -0.46 (0.19) | 0.042* |
| **Default mode network** | 0.04 (0.24) | 0.898 | -0.59 (0.20) | 0.021* | -0.33 (0.18) | 0.116 |
| **Somatomotor** | -0.09 (0.24) | 0.742 | -0.44 (0.19) | 0.058 | -0.30 (0.17) | 0.129 |
| **Salience** | -0.12 (0.24) | 0.720 | -0.35 (0.20) | 0.126 | -0.34 (0.18) | 0.116 |
| **Visual** | -0.19 (0.23) | 0.528 | -0.65 (0.19) | 0.019* | -0.53 (0.17) | 0.021* |

<sup>a</sup>Model adjusted for age, age<sup>2</sup>, sex, scanner type, worst injury of moderate/severe injury severity (versus mild TBI), time since most recent injury, and training/testing set. Each row represents a distinct multiple linear regression model (and models adjusting for lesion run separately); volumetrics were averaged bilaterally and z-standardized to mean 0 and standard deviation 1 for interpretability
\*Statistically significant at 0.05 threshold after adjustment for multiple comparisons using False Discovery Rate (FDR) methods

### Associations of cluster with neuroimaging network-based volumes

We did not observe any demographic differences between participants with and without MRI (Supplementary Table 4). As a sensitivity analysis, we also re-ran the cluster analysis among the subgroup of participants who had MRI data, and observed a largely similar clustering pattern to the full sample (Supplementary Fig 6).

We illustrate raw (unadjusted) mean volumes by cluster in Supplementary Fig 7. In Table 4, we have included a series of multivariable linear regression models for the seven Yeo cortex networks, adjusted for age, age-squared, sex, scanner type, injury chronicity, injury severity, and training/validation set. We found that participants in Cluster 2_cognitive_, compared to Cluster 4_fewest deficits_, had significantly lower volumes in the executive control, dorsal attention, limbic, default mode, and the visual networks. Participants in Cluster 3_mood/behavior_ had significantly lower volumes in dorsal attention, limbic, and visual networks compared to Cluster 4_fewest deficits_. Participants in Cluster 1_mixed_ generally had lower mean volumes than Cluster 4_fewest deficits_, but these findings did not reach statistical significance.

From our linear regression models, we plotted estimated marginal means (adjusted for covariates) for each of the four clusters by cortical network (Fig 5). Cluster 2_cognitive_ had the lowest volumes in every network, particularly default mode and executive control. Both Cluster 2_cognitive_ and Cluster 3_mood/behavior_ had similarly low marginal mean volumes in limbic and visual networks relative to the other two clusters (Cluster 1_mixed_ and Cluster 4_fewest deficits_). In contrast to comparisons with Cluster 4_fewest deficits_, we found in post-hoc pairwise comparisons that there were largely no significant differences in network-specific volumes between the three symptomatic phenotypic comparisons (e.g., Cluster 1_mixed_ and Cluster 2_cognitive_, Cluster 2_cognitive_ and Cluster 3_mood/behavior_) (Supplementary Table 5).

**Fig 5:**
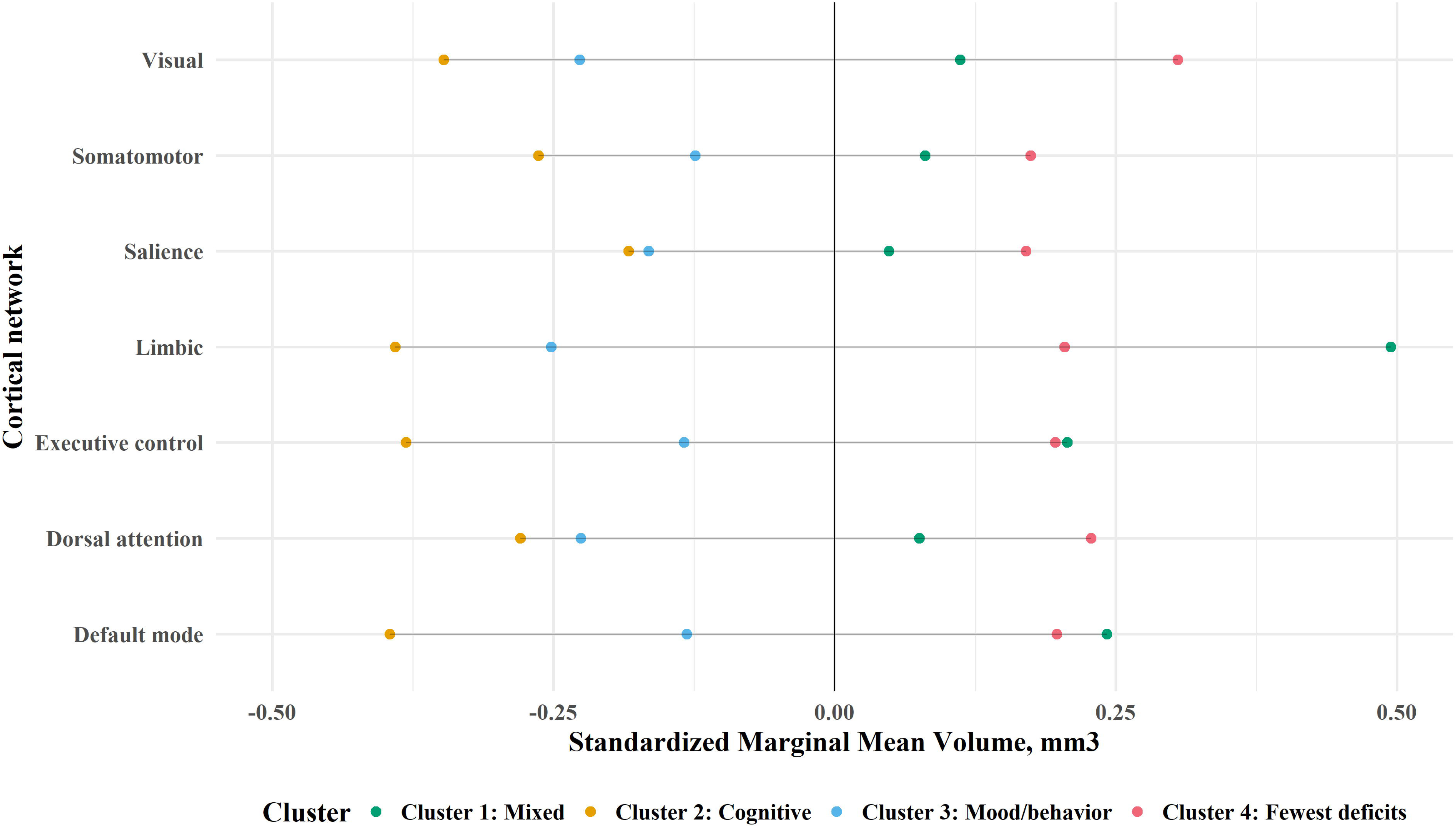
Dot plot of model-based estimated marginal mean cortical volume by cluster. Model-based estimated marginal mean cortical volume (mm^3^) by cluster. The estimated marginal mean corresponds to the least-squares mean at each level of the cluster, adjusting for model covariates (age, age-squared, sex, scanner type, injury severity, injury chronicity, and training set). All volumes were standardized by network to have a mean of 0 and standard deviation of 1 for the sample. Therefore, values below 0 by cluster can be interpreted as below average volumes, and above 0 can be interpreted as above average in the sample.

## Discussion

We identified four clinical clusters, or phenotypes, in a large, well-characterized cohort of individuals with chronic TBI. The phenotypes were associated with network-based measures of cortical volume, providing a putative neuroanatomic basis for the observed phenotypes.

Identification of four distinct phenotypes, which included 1) mixed deficits, 2) predominant cognitive deficits, 3) predominant mood/behavioral deficits, and 4) relatively few deficits, was facilitated by the breadth of clinical data in LETBI study^27^ across domains of cognition, mood, behavior, physical and motor function, as well as the long-term follow up that extends beyond one year post injury. Collectively, these observations suggest that machine learning consolidation of high-dimensional, multimodal clinical data may be used to identify chronic TBI phenotypes that are grounded in the underlying pathophysiologic mechanisms of symptom development in chronic TBI. The use of more granular clinical classification that is based on domain-specific deficits with pathophysiological relevance, as opposed to crude indices of disability, may prove valuable to inform patient stratification, clinical trial inclusion, and outcome measurement.^63^

In the current study, the greatest proportion of participants (40.0%) fit into Cluster 4_fewest deficits_, which includes those with the fewest persistent cognitive, mood, neurobehavioral, and physical deficits relative to the current sample of individuals with chronic TBI. This finding illustrates the heterogeneity of chronic TBI deficits, while also emphasizing that many long-term survivors of head trauma have little or no longstanding disability, and the progressive decline observed in some survivors^64,65^ is far from universal. However, the remainder of the sample (60%) are living with varying degrees of impairment across domains. More than a quarter of the sample (27.7%) had predominant mood/behavioral deficits, while 20.5% had predominant multi-domain cognitive impairment. The smallest phenotype (11.8% of the sample) was characterized by mixed deficits spanning multiple neurobehavioral, cognitive, motor, and general health measures.

Of all the studies included in a recent review of TBI phenotyping efforts,^20^ only two collected cognitive, mood, and behavioral data from adults with moderate-severe TBI who were assessed more than one-year post-injury. Juengst et al.^66^ identified four groups characterized by poor mood and behavioral function without cognitive impairment (similar to Cluster 3_mood/behavior_ in the current study), good mood and average behavior with mildly impaired-intact cognitive function (most similar to our Cluster 4_fewest deficits_); high anxiety, poor behavioral function, and relatively intact cognitive performance (perhaps most similar to our Cluster 1_mixed_); and good emotional function with behavioral impairment and severe cognitive impairment (similar to our Cluster 2_cognitive_). Sherer and colleagues^67^ used similar cluster analysis methods to the present study across 12 different measures of TBI recovery, and observed five different clusters that align reasonably well with those presented herein; measures of self-awareness, social support, and performance validity split the group that most similar to the current Cluster 3_mood/behavior_ into two groups. Despite the use of different assessment batteries and constructs measured, the overlap in chronic TBI phenotypes identified across studies suggests that there are common clinical profiles. Notably, neither of these two chronic TBI phenotyping studies included biomarker data. Therefore, the current work provides an important extension by examining neuroanatomic differences across phenotypes.

To elucidate the neuroanatomic basis for TBI outcome phenotypes, we tested for network-based imaging correlates of the observed clinical phenotypes. In the Cluster 2_cognitive_ phenotype, we found significantly lower average cortical volumes in executive control, dorsal attention, limbic, default mode and visual networks, relative to the group with the fewest deficits. These observations are consistent with extensive prior work demonstrating that the structure and function of these networks, particularly the executive control and default mode networks, are associated with deficits in higher-order cognition. Indeed, the pathophysiologic link between brain network injury and clinical symptoms in individuals with TBI is well established.^68^ We build upon prior foundational work by showing that a network-based cortical volumetric measure derived from clinically accessible T1-weighted MRI data has the potential to serve as an affirmatory biomarker indicative of an organic, neuroanatomical basis for cognitive symptomatology after TBI. Notably, the Cluster 2_cognitive_ phenotype had the lowest educational attainment relative to all other phenotypes. Lower education in this phenotype could reflect lower cognitive reserve, which could be an additional contributor to lower observed cognitive performance in this subgroup.

In the Cluster 3_mood/behavior_ phenotype, we found lower volumes in dorsal attention, limbic, and visual networks as compared to the phenotype with the fewest deficits. This observation aligns with research implicating pre-frontal-limbic^69,70^ and attentional^71^ network circuitry in the pathophysiology of depression and mood.^72^ Another functional MRI study^73^ demonstrated abnormal connectivity in the visual-attentional network of patients with depression relative to controls. While our data indicate that decreases in dorsal attention, limbic, and visual network are sensitive biomarkers distinguishing the mood/behavioral and fewest deficit phenotypes, in our post-hoc pairwise analysis, we did not detect significant mean differences in cortical network volumes between Cluster 2_cognitive and_ Cluster 3_mood/behavior_ phenotypes. This suggests that cortical network volumes may not be sufficiently *specific* to differentiate between cognitive and mood/behavioral phenotypes. Specifically, the cortical network volumes that are reduced in the predominant mood/behavior cluster (i.e., dorsal attention, limbic, visual networks) are also diminished in individuals with the cognitive phenotype.

The observed network-based cortical volumes in the Cluster 1_mixed_ phenotype were contrary to our expectation based this group’s broad clinical presentation. In fact, the approximately 11% of individuals in this phenotype had marginal mean cortical volumes closer to Cluster 4_fewest deficits_ versus the other two symptomatic phenotypes (Cluster 2_cognitive_ or Cluster 3_mood/behavior_). There are several potential explanations for our findings. First, the observation that individuals in Cluster 1_mixed_ had a range deficits does not necessarily equate to all of those symptoms being severe across the board. This is apparent in their performance-based cognitive test scores; the Cluster 2_cognitive_ phenotype had noticeably worse cognitive impairment across most neuropsychological tests relative to Cluster 1_mixed_. It is also possible the cortical network-based biomarkers we evaluated in this study were not sufficiently sensitive to detect a signal in this small subgroup (n=36) with mixed trait deficits that lack a defining clinical feature. Future studies might focus instead on diffusion-based neuroimaging biomarkers (i.e., mean diffusivity), or fluid biomarkers that are thought to better track to TBI polypathology (e.g., glial fibrillary acidic protein^74^ or neurofilament light^75,76^). The phenotype with mixed trait clinical symptoms without distinguishable pathology could also represent a more vulnerable subgroup, potentially driven by an unmeasured confounder (i.e., low socioeconomic status, early life or contemporaneous life adversity) that leads to a lower threshold to exhibit symptoms than would be predicted by their neuroimaging biomarkers. Studies using residual statistical methods have described this phenomena when quantifying discrepancies between observed and predicted cognitive ability as a function of brain pathology.^77^

Given the wide scope of clinical domains assessed, the current study permitted some hypothesis-generating observations with respect to patterns of co-occurrence of impairment across and within phenotypes that are not pronounced in our high-level qualitative phenotype labels. For example, motor impairment was greatest in the mixed trait phenotype (Cluster 1_mixed_), followed by the predominant cognitive impairment phenotype (Cluster 2_cognitive_). Those with the least cognitive impairment (Cluster 3_mood/behavior_ and Cluster 4_fewest deficits_) did report the highest rates of illicit drug and alcohol use, respectively, which is consistent with prior findings suggesting some health risk behaviors may be greatest among the most high-functioning TBI survivors who have the level of independence required to access substances. ^78,79^

The role of age and injury chronicity are important to contextualize our findings for the Cluster 4_fewest deficits_ phenotype. This group was on average ∼10 years older than all other phenotypes, and were the furthest removed from their most recent TBI. While counterintuitive based on evidence of an interaction between older age and injury chronicity in TBI^80^, our findings could be explained by age-norming of neuropsychological data. For example, performance on a cognitive test for a 70 year old participant in the LETBI study was effectively compared to a similar 70 year old in the general population. Observed TBI-related differences in cognitive performance will be more pronounced at younger ages, where the referent general population exhibits fewer baseline cognitive deficits on average. Furthermore, when we visually compared unadjusted differences in volume by network (Supplementary Fig 7), we saw modest differences in volume by phenotype. However, when we adjusted our regression models for age and age-squared (along with other covariates), we detected significant differences in volume across multiple networks between phenotypes. Our findings underscore the importance of considering linear and non-linear representations of age as a confounder when interpreting normative-based scoring of cognitive tests and associated objective brain pathology in chronic TBI studies.^60^

### Limitations and Strengths

There are limitations of the present study. Our data are cross-sectional in nature, and we cannot make conclusions on decline over time of clinical phenotypes, nor can we imply evidence of post-traumatic neurodegeneration from our imaging biomarkers. The LETBI study involves a 4-6 hour study visit consisting of comprehensive clinical and neuropsychological testing, in addition to MRI assessment, which may have led to a selection bias in favor of higher functioning TBI survivors. This phenomenon may have restricted enrollment of more impaired older adults with TBI, such that roughly two-thirds of enrolled participants in our study over age 65 were of the phenotype with fewest deficits. Our sample represented participants across the adult lifespan, though it did tend to skew older and more educated compared to other multicenter TBI studies.^81,82^ We acknowledge that our MRI sample was smaller than the clinical phenotype analysis sample. Some participants did not receive a research MRI due to personal choice (i.e., claustrophobia) or contraindications like metal in their body or pregnancy. However, since we did not observe any demographic differences between those who did and did not receive MRI (see Supplementary Table 4), and because our cluster results from Aim 1 were largely replicated in the subgroup who received MRI (see Supplementary Fig 6), we do not have reason to believe this led to selection bias in our findings. While our current study of n=281 individuals is among the largest multimodal studies of its kind ever conducted in a chronic TBI sample, we determined a four-cluster solution was the optimal fit in our sample. Because of our classification schema, we were unable to dig deeper into specific domains within cognition (i.e., attention, memory) and mood/behavior (i.e., depression, impulsivity), and their associated neuroanatomic signatures. This is a future direction with the accrual of larger samples with more power to detect a greater number of clusters.

Major strengths of our study were the inclusion of individuals spanning a wide range of TBI severities and chronic outcomes, which is particularly important given that most prior studies attempting to classify clinical phenotypes have exclusively focused on the acute period in the first year after mild TBI. Finally, our pre-processing steps for the clinical and MRI data overcome potentially major sources of selection bias from past work. We imputed selective missing data in our clinical data, which allowed us to avoid excluding the individuals who did not have complete clinical data across 41 measures. We also developed an advanced processing method to perform cortical volumetric measurements in individuals with large lesions, whereas many prior TBI neuroimaging studies systematically excluded this non-random subgroup of individuals.^46^

### Conclusions

The present study used multimodal data from the LETBI cohort to identify four distinct clinical phenotypes after TBI. We found that individuals with predominant cognitive deficits and mood and behavioral deficits had the largest differences in network-specific cortical volumes – particularly in the default mode, executive control, limbic and visual networks – compared to individuals with relatively few deficits clinical outcomes. If these findings are replicated in future studies, they may inform future chronic classification ontologies and clinical trial stratification methods to facilitate the development and validation of personalized treatments for patients living with chronic TBI. Future studies should build upon this work by investigating the stability of observed phenotypes in other chronic TBI cohorts, and in longitudinal multimodal datasets that include diverse imaging and fluid biomarkers.

## Data Availability

Data are available through request from the Federal Interagency Traumatic Brain Injury Research (FIT-BIR) program.

https://fitbir.nih.gov/

## Abbreviations

(TBI): Traumatic brain injury
(LETBI): Late Effects of TBI Study

## Acknowledgements

Casey Cheng developed the PCA theoretical depiction used in Fig 2, and it is used in this figure with his permission.

## Supplementary material

Supplementary material is available at *Brain* online.

## Data availability

The data used to generate this study is available upon request from the Federal Interagency Traumatic Brain Injury Research (FITBIR) platform (https://fitbir.nih.gov/content/access-data).

