## Supplementary Table 1 for "Distinct clinical phenotypes and their neuroanatomic correlates in chronic traumatic brain injury"

### **Supplementary Table 1:** Details on lesion correction ratings.

| RATING | ASEG | SURFACE |
| --- | --- | --- |
| 1 | Cortical GM/WM segmentation is free of minor errors, subcortical and cerebellar contrast is not oversaturated, subcortical and cerebellar segmentations are free of minor errors and deviations | Pial and white surfaces closely follow anatomical boundaries, no minor areas excluded from either |
| 2 | Minor deviations observable in GM/WM segmentations, contrast in cerebellar and subcortical areas may be difficult to assess, cerebellar and subcortical segmentations appear reasonable with some over and under-labeling. Requires minor manual edits | Pial and white surfaces may exclude some parts of the cortex, but no major exclusions are observable. Requires minor manual edits |
| 3 | Obvious deviations observable in GM/WM segmentations, slightly worse than rating '2' and requiring major manual edits | Obvious exclusions on the surfaces requiring major manual edits, but not enough to warrant exlusion |
| 4 | EXCLUDE - Major deviations observable in GM/WM segmentations, little to no contrast in subcortical and cerebellar areas, major deviations in subcortical and cerebellar labels | EXCLUDE - Major disruptions in pial and white surfaces, excludes and/or mislabels areas of cortex |
