## Supplementary Table 2 for "Distinct clinical phenotypes and their neuroanatomic correlates in chronic traumatic brain injury"

### **Supplementary Table 2:** Rank Order of Top 12 Contributors and Loadings of Individual Measures Making up Principal Components (PC1-PC3)

| Rank order | PC1 Measures (24.8% variance) | PC1 loadings | PC2 Measures (14.5% variance) | PC2 loadings | PC3 Measures (6.0% variance) | PC3 loadings |
| --- | --- | --- | --- | --- | --- | --- |
| 1 | RAND-Social Function | 0.727 | CVLT-SDFR | 0.692 | UPDRS-Part 3 Total | 0.609 |
| 2 | RAND-Emotional Wellbeing | 0.693 | CVLT-Immediate Recall | 0.688 | RAND Physical Function | -0.486 |
| 3 | RAND-Energy/Fatigue | 0.670 | CVLT-LDFR | 0.677 | Sit-to-stand | 0.456 |
| 4 | QOL-Social | 0.669 | WMS-LM2 | 0.590 | RAND-Emotional Limitations | 0.405 |
| 5 | SWLS | 0.668 | WMS-LM1 | 0.551 | CVLT-Semantic Clustering | 0.358 |
| 6 | QOL-Anxiety | -0.667 | QOL-Fatigue | 0.499 | BIS-Motor | -0.299 |
| 7 | QOL-Fatigue | -0.644 | RAND-Energy/Fatigue | -0.493 | Trails A | 0.289 |
| 8 | QOL-Depression | -0.641 | REY-Delay Recall | 0.469 | Assist-Illicit Substance | -0.283 |
| 9 | WAIS Coding | 0.635 | REY-Immediate Recall | 0.450 | Grip strength-Dominant Hand | -0.283 |
| 10 | MIDUS Health | -0.628 | Symbol search | 0.440 | CVLT-SDFR | 0.274 |
| 11 | RAND-General Health | 0.606 | RAND-General Health | -0.433 | Assist-Alcohol | -0.270 |
| 12 | Symbol search | 0.599 | COWAT-Animals | 0.430 | RAND-Pain | -0.262 |
