## Supplementary Table 3 for "Distinct clinical phenotypes and their neuroanatomic correlates in chronic traumatic brain injury"

### **Supplementary Table 3:** Cluster Validity Indices

|  | Connectivity | Dunn Index | Silhouette |
| --- | --- | --- | --- |
| 3 group | 25.765 | 0.095 | 0.328 |
| 4 group | 39.06 | 0.099 | 0.320 |
| Optimal number of clusters based on given index | 3 group | 4 group | 3 group |
