## Supplementary Table 4 for "Distinct clinical phenotypes and their neuroanatomic correlates in chronic traumatic brain injury"

### **Supplementary Table 4:** Characteristics of the sample with and without MRI

|  | Without MRI (n=99) | With MRI (n=182) | p-value |
| --- | --- | --- | --- |
| Age, Mean (SD) | 56.2 (17.6) | 58.3 (15.0) | 0.296 |
| Age group, n (col %)  <40  40-54  55-64  65+ | 19 (19.2%)  27 (27.3%)  20 (20.2%)  33 (33.3%) | 21 (11.5%)  51 (28.0%)  47 (25.8%)  63 (34.6%) | 0.317 |
| Sex, Female (col %) | 38 (38.4%) | 58 (31.9%) | 0.271 |
| Education, n (col %)  <College  ≥College degree | 23 (23.2%)  76 (76.8%) | 54 (29.7%)  128 (70.3%) | 0.248 |
| Race, n (%)  White  Black  Other | 82 (82.8%) 8 (8.1%) 9 (9.1%) | 160 (87.9%) 11 (6.0%)  11 (6.0%) | 0.491 |
| Hispanic ethnicity, n (col %) | 7 (7.1%) | 16 (8.8%) | 0.615 |
| Marital status, n (col %)  Never married  Married/partnered  Divorced/widowed | 29 (29.3%)  45 (45.5%)  25 (25.3%) | 44 (24.2%)  76 (41.8%) 62 (34.1%) | 0.293 |
| Employment, n (col %)  Working/Student  Unemployed  Retired  Disabled  Other | 33 (33.3%)  8 (8.1%)  27 (27.3%)  28 (28.3%)  3 (3.0%) | 60 (33.0%)  10 (5.5%)  58 (31.9%)  40 (22.0%) 14 (7.7%) | 0.345 |
