## Supplementary Table 5 for "Distinct clinical phenotypes and their neuroanatomic correlates in chronic traumatic brain injury"

**Supplementary Table 5:** Pairwise comparisons of least squares mean difference in cortical network volume by cluster^¥^

| **Pairwise comparison** | **Network** | **Least Squares** **Mean Difference (95% CI)** | **p-value** |
| --- | --- | --- | --- |
| **Cluster 1_mixed_ – Cluster 2_cognitive_** | Executive control | 0.59 (-0.08, 1.25) | 0.103 |
|  | Dorsal attention | 0.35 (-0.30, 1.01) | 0.495 |
|  | Limbic | 0.89 (0.17, 1.60) | 0.008* |
|  | Default mode network | 0.64 (-0.04, 1.31) | 0.073 |
|  | Somatomotor | 0.34 (-0.31, 1.00) | 0.525 |
|  | Salience | 0.23 (-0.44, 0.90) | 0.808 |
|  | Visual | 0.46 (-0.18, 1.10) | 0.245 |
| **Cluster 1_mixed_ – Cluster 3_mood/behavior_** | **Network** | **Least Squares** **Mean Difference (95% CI)** | **p-value** |
|  | Executive control | 0.34 (-0.28, 0.96) | 0.483 |
|  | Dorsal attention | 0.30 (-0.31, 0.91) | 0.575 |
|  | Limbic | 0.75 (0.08, 1.41) | 0.021* |
|  | Default mode network | 0.37 (-0.26, 1.00) | 0.417 |
|  | Somatomotor | 0.20 (-0.41, 0.82) | 0.821 |
|  | Salience | 0.21 (-0.41, 0.84) | 0.812 |
|  | Visual | 0.34 (-0.26, 0.93) | 0.452 |
| **Cluster 2_cognitive_ – Cluster 3_mood/behavior_** | **Network** | **Least Squares** **Mean Difference (95% CI)** | **p-value** |
|  | Executive control | -0.25 (-0.76, 0.25) | 0.593 |
|  | Dorsal attention | -0.05 (-0.56, 0.45) | 0.993 |
|  | Limbic | -0.14 (-0.69, 0.41) | 0.914 |
|  | Default mode network | -0.26 (-0.79, 0.26) | 0.555 |
|  | Somatomotor | -0.14 (-0.65, 0.37) | 0.891 |
|  | Salience | -0.02 (-0.54, 0.50) | 0.959 |
|  | Visual | -0.12 (-0.61, 0.37) | 0.919 |

^¥^: Pairwise comparisons with Cluster 4 (fewest deficits) not included in this table because it is included in Table 3
