## Supplementary Fig 1 for "Distinct clinical phenotypes and their neuroanatomic correlates in chronic traumatic brain injury"

### **Supplementary Fig 1:** Exemplar of MRI scan before and after implementation of novel lesion correction methodology

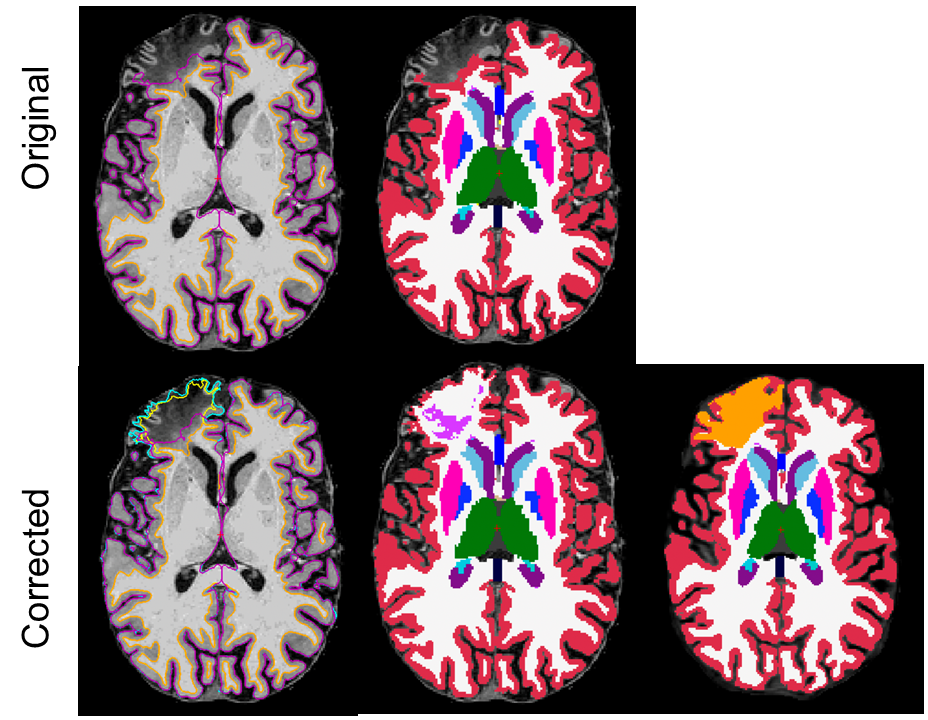

Supplementary Fig 1 caption: The “before” and “after” MRI scans illustrating the implementation of the novel lesion correction methodology.
