## Supplementary Fig 2 for "Distinct clinical phenotypes and their neuroanatomic correlates in chronic traumatic brain injury"

### **Supplementary Fig 2:** Initial cutpoint selected by the Hierarchical Cluster Analysis algorithm

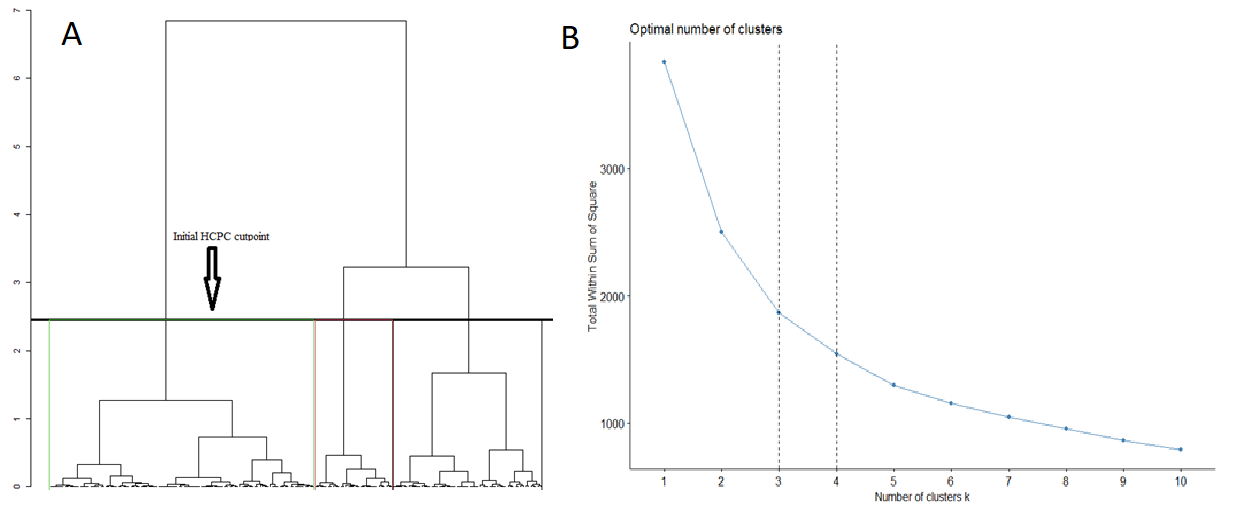

Supplementary Fig 2 caption: 1A) The Hierarchical Cluster Analysis on Principal Components (HCPC) algorithm outputs an initial cutpoint shown here. The algorithm suggested 3 components, which we tested empirically to assess reliability and validity versus other specifications. 1B) The total within cluster sum of squares, a measure of variance within cluster. There was an elbow at 3 and 4 cluster specifications, which were further empirically compared.
