## Supplementary Fig 3 for "Distinct clinical phenotypes and their neuroanatomic correlates in chronic traumatic brain injury"

**Supplemental Fig 3**: Hierarchical cluster group assignment (validation set)

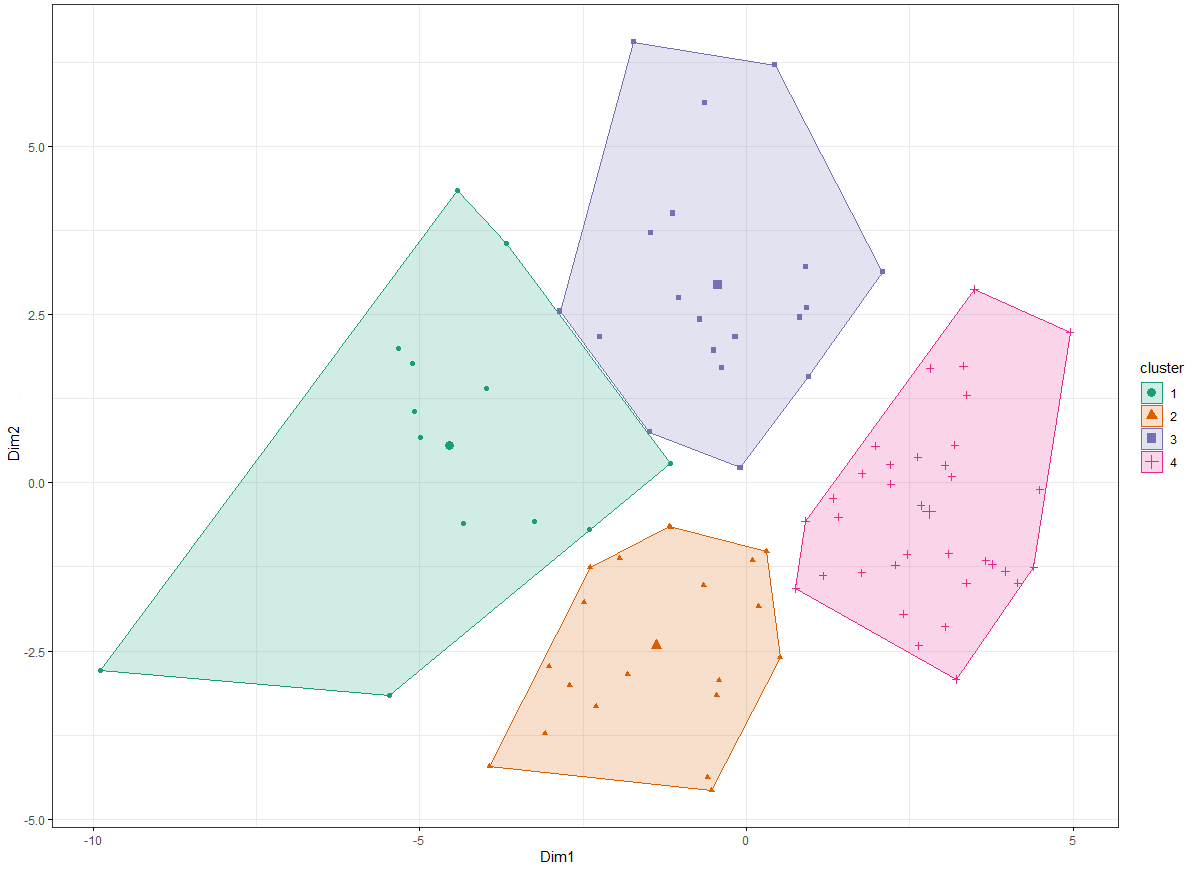

Supplemental Fig 3 caption: Hierarchical cluster group assignment in the validation set. The results are based on a HCPC analysis. Here, each participant in the validation sample is depicted in the x-y coordinate space based on their PC1 vs. PC2 scores. The cluster membership of each participant is color coded.
