## Supplementary Fig 4 for "Distinct clinical phenotypes and their neuroanatomic correlates in chronic traumatic brain injury"

### **Supplementary Fig 4:** Three group cluster classification in training (Panel A) and validation (Panel B) sets

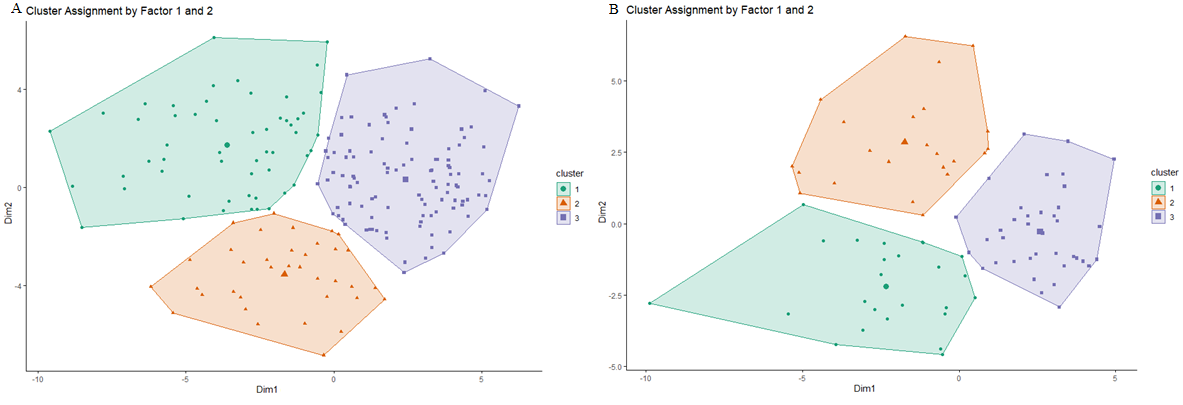

Supplementary Fig 4 caption: The three cluster specification in the training (panel A) and validation (panel B) sets. There was poor reliability of cluster centroids in the internal validation versus the training sets; therefore, the three cluster specification was deemed to have unacceptable reliability.
