## Supplementary Fig 5 for "Distinct clinical phenotypes and their neuroanatomic correlates in chronic traumatic brain injury"

**Supplemental Fig 5**: Heat map characterizing average values of neurobehavioral measures by cluster assignment (validation set; n=86)

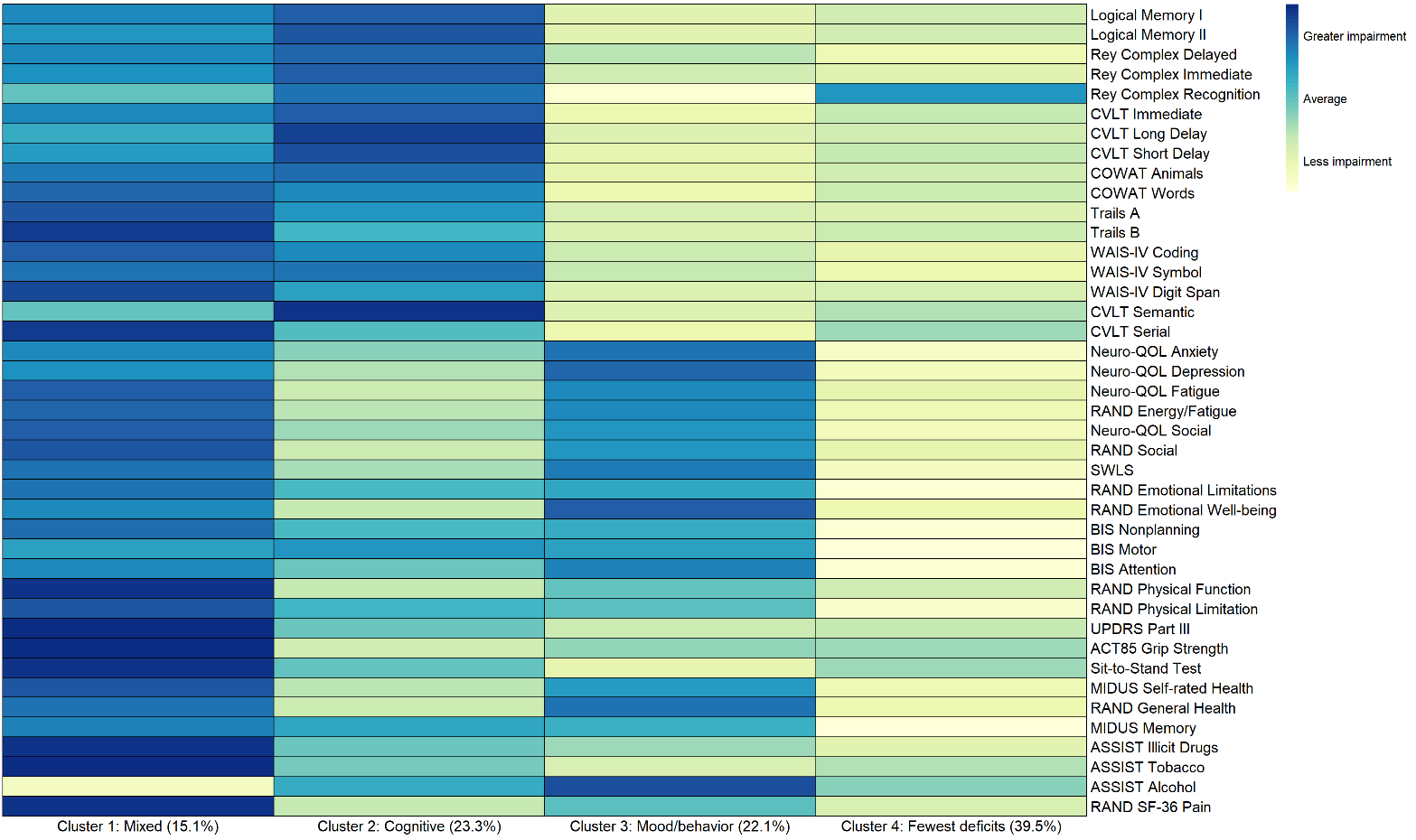

Supplemental Fig 5 caption: Heat map characterizing average values of neurobehavioral measures by cluster assignment in the validation set. The measures were transformed such that darker colors represent greater impairment, and lighter colors represent less impairment. The qualitative descriptors are consistent as we observed in the training set: Cluster 1: Mixed trait deficits; Cluster 2) Predominant cognitive deficits; Cluster 3: Predominant mood and behavioral deficits; Cluster 4: Relatively few deficits.
