## Supplementary Fig 6 for "Distinct clinical phenotypes and their neuroanatomic correlates in chronic traumatic brain injury"

**Supplementary Fig 6:** Sensitivity analysis: replication of cluster analysis among only participants who have an MRI

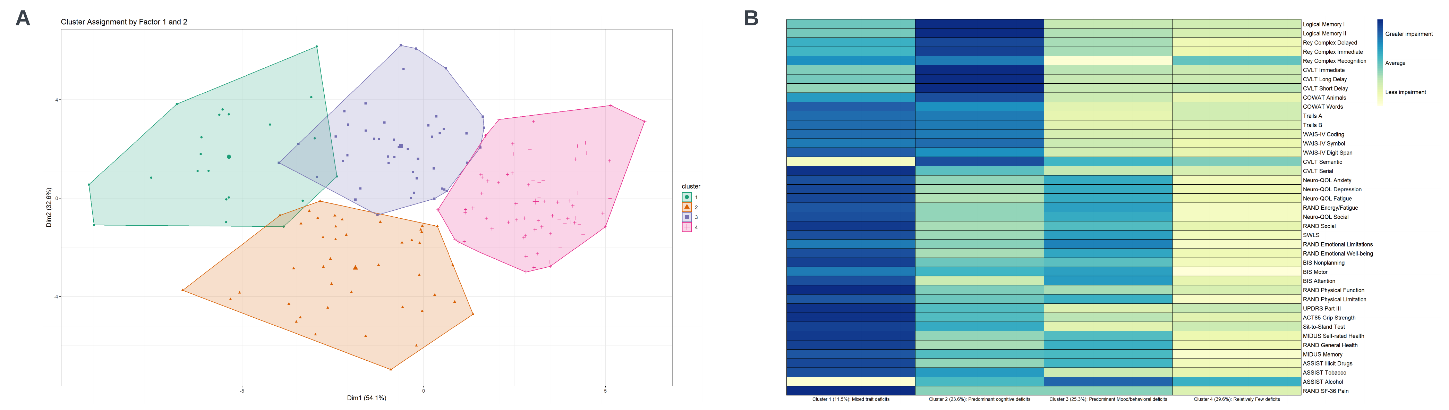

Supplementary Fig 6 caption: Panel A is the four group cluster grouping as specified in Aim. Analyses were run only among participants with MRI to assess the reliability of Aim 1 results among this subgroup to determine if results are generalizable. Panel B is the heat map based on the cluster groupings in Panel A among the subgroup with MRI. We determined the results are largely generalizable to the findings from Aim 1.
