## Supplementary Fig 7 for "Distinct clinical phenotypes and their neuroanatomic correlates in chronic traumatic brain injury"

### **Supplementary Fig 7:** Raw (unadjusted) mean volumes by cluster and network

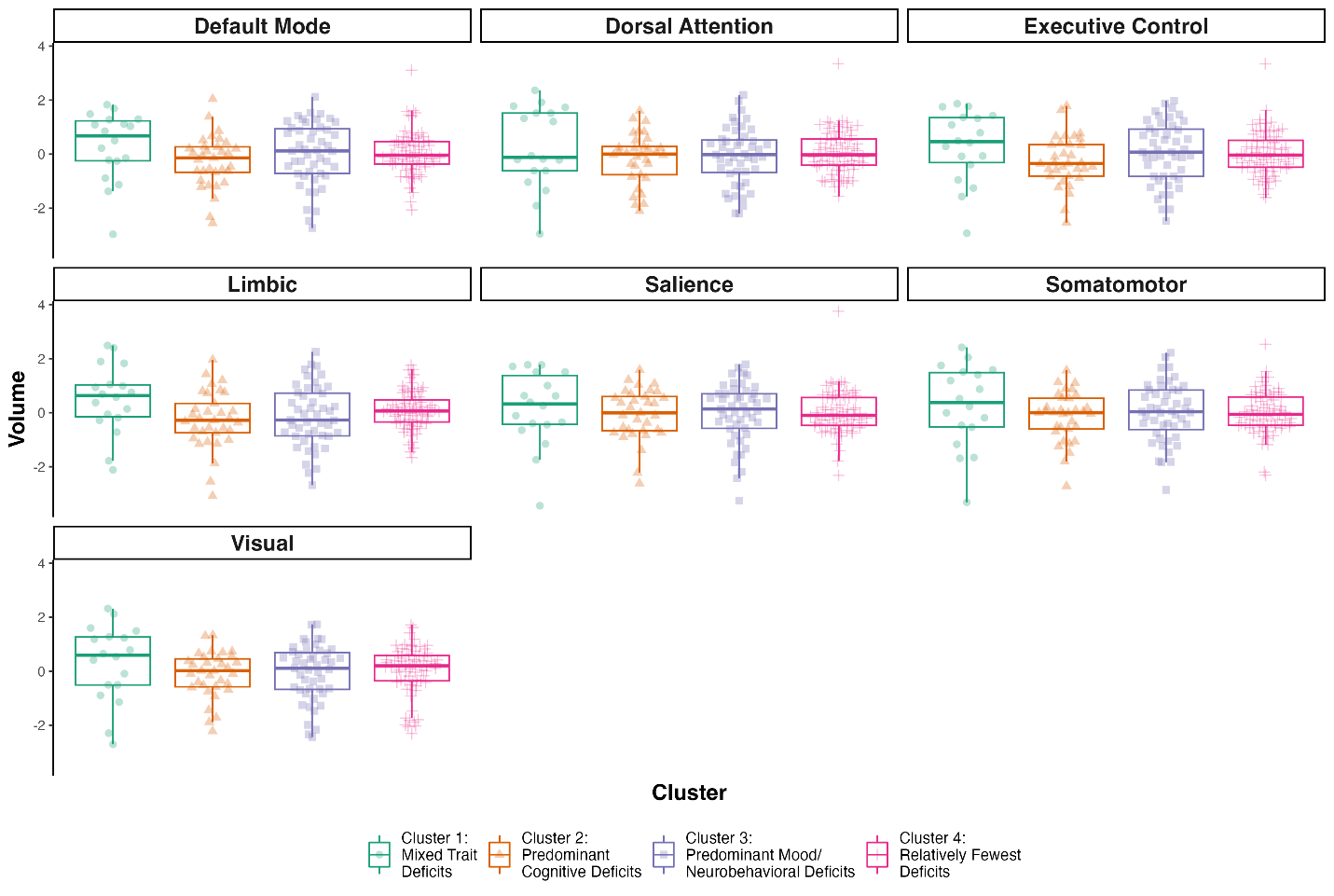

Supplementary Fig 7 caption: Unadjusted (raw) mean network volumes for each of the 7 networks from the Yeo-7 atlas by phenotype group.
